# Unstable sleep and rest-activity rhythms in adolescents at-risk for bipolar disorder: links to mood symptoms and the effect of sleep stabilization

**DOI:** 10.64898/2025.12.29.25342858

**Authors:** João Paulo Lima Santos, Lauren Keller, Amy G. Hartman, Simey Chan, Mary L. Phillips, Adriane M. Soehner

## Abstract

**Background:** Adolescents with a parental history of bipolar disorder (BD; High-Risk) are more likely to develop BD than those without such history (Low-Risk). Actigraphy studies in adults indicate that instability in sleep and rest-activity rhythms (RAR) may be associated with progression to BD. However, it remains unclear whether High-and Low-Risk adolescents are differentially affected by sleep/RAR instability and their impact on emerging mood symptoms.

**Methods:** We recruited 50 adolescents unaffected by BD (mean age[SD]=15.97[1.31], 58% female) stratified into High-Risk (N=22) and Low-Risk (N=28) groups. All participants completed two weeks of Baseline actigraphy; a subset of 11 High-Risk participants then completed a two-week sleep Stabilization Manipulation at home. Depression and mania rating scales were collected at Baseline and post-Manipulation. Covariates included age, sex at birth, total tracking days, and ratio of weekday to weekend tracking days; false-discovery rate correction was applied.

**Results:** At Baseline, sleep/RAR instability did not differ between High and Low-Risk groups (P-values>0.05). Among High-Risk participants only, greater sleep duration variability was linked to higher levels of depressive (β[95%CI]=0.012[0.006,0.018]) and mania (β[95%CI]=0.011[0.003,0.019]) symptoms, and lower Circadian Function Index (CFI; composite measure of circadian activity robustness) was linked to greater mania symptoms (β[95%CI]=-8.236[-12.525,-3.940]). The Stabilization protocol reduced variability in sleep duration (P<0.001), midsleep (P=0.005), and CFI (P=0.044) in High-Risk adolescents, and reduced subthreshold mania symptoms, but this did not survive multiple comparison correction.

**Conclusions:** These findings highlight that sleep/RAR instability may have a stronger impact on mood in High-Risk adolescents but simple behavioral interventions stabilizing sleep and rhythms could buffer against BD risk.

## 1. INTRODUCTION

For many individuals living with Bipolar disorder (BD), the condition follows a severe, chronic, and debilitating course(Kessler et al., 2005; Merikangas et al., 2007). BD often emerges during adolescence and young adulthood(Kessler et al., 2005; Merikangas et al., 2007), but diagnosis and treatment are frequently delayed due to atypical clinical presentations and socioeconomic barriers(Boström et al., 2024; Post et al., 2020). Individuals with a parental history of BD (referred to as High-Risk) are ten times more likely to develop the disorder than those without such history (Low-Risk)(Birmaher et al., 2009; Kessler et al., 2005). This heightened risk in adolescents is particularly alarming, as an earlier onset of BD is associated with more long-term issues, including more severe illness progression(Mignogna and Goes, 2024), higher rates of co-occurring psychiatric conditions(Yapıcı Eser et al., 2020), and detrimental changes to the brain(Lima Santos et al., 2022). Given that family history of BD is a well-established static risk marker, it is critical to establish modifiable, causal risk factors in at-risk adolescents to reduce the likelihood of BD and its associated adverse outcomes(Kraemer et al., 1997). One such factor is instability of sleep-wake patterns and rest-activity rhythms (RAR), which may exacerbate neurobehavioral vulnerabilities in individuals with or at risk for BD and accelerate illness progression(Ng et al., 2015).

Among individuals with BD, sleep/RAR instability is present in acute depressive and mania mood episodes and persists in the inter-episode period, suggesting it may be a behavioral indicator of biological vulnerability for BD(Alloy et al., 2017; Bullock and Murray, 2014; Ng et al., 2015). Additionally, escalating sleep/RAR instability can precede new mood episodes(Melo et al., 2016; Ortiz et al., 2025), serving as an early warning sign of recurrence. However, the findings regarding sleep/RAR instability in High-Risk adolescents are mixed(Faria et al., 2015; Melo et al., 2016; Ng et al., 2015; Scott et al., 2022). While some studies report greater instability of sleep/RAR outcomes(Faria et al., 2015; Ng et al., 2015), others report no consistent or significant differences between High-and Low-Risk groups(Melo et al., 2016; Scott et al., 2022). In adolescence, a confluence of biopsychosocial factors contributes to a marked rise in sleep/RAR instability, which coincides with escalating risk for BD(Birmaher et al., 2009; Carskadon, 2011; Cole et al., 1992; Duffy, 2014; Frank et al., 2015; Kessler et al., 2005). Due to neurodevelopmental differences among High-Risk adolescents(Deveney et al., 2012; Kim et al., 2012; Manelis et al., 2016; Singh et al., 2014), it is possible that even a normative shift in sleep/RAR instability could confer more adverse effects on mood than typically developing peers. Thus, even in the absence of notable case-control differences in sleep/RAR instability among High vs Low-Risk adolescents, sleep/RAR instability could be more strongly coupled with mood disruption among High-Risk adolescents. If this notion is supported, it would bolster evidence for early interventions targeting sleep/RARs to prevent symptom escalation and, potentially, delay BD onset.

Studies have begun to evaluate whether stabilizing sleep and RARs can reduce symptom burden and improve outcomes in individuals with BD(Harvey et al., 2015; Simjanoski et al., 2023). For instance, one pilot clinical trial evaluated a BD-specific adaptation of cognitive behavioral therapy for insomnia (CBTI-BP(Harvey et al., 2015)) versus psychoeducation, finding that adults with BD type I who received the active treatment experienced fewer relapses of hypomania/mania and fewer days spent in mood episodes compared to those in a psychoeducation group(Harvey et al., 2015). Additionally, Interpersonal and Social Rhythm Therapy (IPSRT(Goldstein et al., 2014a; Hlastala et al., 2010)) – a component of which focuses on stabilizing daily routines and sleep schedules – has been evaluated as an early intervention in a couple small pilot trials among adolescents with or at High-Risk for bipolar spectrum disorders. IPSRT reported similar outcomes to CBTI-BP, including reduction of the severity and/or duration of mania symptoms following treatment(Goldstein et al., 2014a; Hlastala et al., 2010). Yet, these multi-component intervention trials did not evaluate improvement in sleep/RAR stability as a specific driver of improvement in symptom status. A brief, therapeutic sleep manipulation may be one means of bridging this empirical gap among High-Risk adolescents.

The goal of this study is to examine the degree to which sleep/RAR instability contributes to mood symptoms in High-Risk adolescents (14-19 years) and to assess the impact of a brief, at-home sleep stabilization manipulation protocol. The first aim was to examine whether sleep/RAR instability differed between High-Risk and Low-Risk adolescents, and whether risk status moderated associations with symptoms of depression and mania. We hypothesized that case-control differences would not be present at this developmental stage, but that sleep and RAR instability would be more strongly associated with mood symptoms in the High-Risk group. Herein, we piloted a 2-week at-home sleep/RAR stabilization protocol, distilled from components of CBT-BP, and explored the degree to which this therapeutic sleep manipulation stabilized sleep/RARs and improved mood symptoms in High-Risk adolescents. We hypothesized that High-Risk adolescents undergoing the stabilization manipulation protocol would show improvements in sleep/RAR instability compared to baseline and explored whether interindividual differences in sleep/RAR changes correlated with changes in self-rated mood.

## 2. MATERIALS AND METHODS

### 2.1. Participants

The present report is based on data from a sleep and neuroimaging pilot study approved by the University of Pittsburgh Human Research Protection Office (STUDY19050314). The current analyses were not preregistered.

Our analytic sample was comprised of adolescents recruited for a sleep and neuroimaging pilot study that were typically developing healthy controls (Low-Risk; N=28) or at elevated risk for bipolar disorder based on parental history of the diagnosis (High-Risk; N=22). Participant characteristics are reported in **Table 1**. All participants were a) 14.0-19.11 years old; b) enrolled during the high school academic year; c) right-handed; d) MRI eligible (i.e., no ferromagnetic metal in the body), e) free of major systemic medical illnesses, neurological disorders, history of head trauma, and intellectual disabilities (parent report); f) free of organic sleep disorders (e.g., apnea, narcolepsy, restless legs syndrome, REM behavior disorder) per a locally-developed semi-structured interview, and g) not pregnant or breastfeeding (self-report and/or urine test).

**Table 1.** Demographic, clinical, and sleep characteristics in the full sample and by group (N=50).

| Variable | Full Sample<br>(N=50) | High-Risk<br>(N=22) <sup>1</sup> | Low-Risk<br>(N=28) | Between-group<br>Statistics <sup>2</sup> |  |
| --- | --- | --- | --- | --- | --- |
| Demographics |  |  |  |  |  |
| Age (Yr; mean[SD]) | 15.97 [1.31] | 15.87 [1.33] | 16.05 [1.31] | t=0.46 | 0.645 |
| Sex at birth (% Female) | 29 (58.00%) | 15 (68.18%) | 14 (50.00%) | χ<sup>2</sup>=1.01 | 0.315 |
| Ethnicity (% Non-Hispanic) | 46 (92.00%) | 21 (95.45%) | 25 (89.29%) | χ<sup>2</sup>=0.07 | 0.785 |
| Race |  |  |  |  |  |
| White (%) | 35 (70.00%) | 15 (68.18%) | 20 (71.43%) | χ<sup>2</sup>=0.09 | 0.958 |
| Asian (%) | 0 (0.00%) | 0 (0.00%) | 0 (0.00%) |  |  |
| Black (%) | 11 (22.00%) | 5 (22.73%) | 6 (21.43%) |  |  |
| Multiple (%) | 4 (8.00%) | 2 (9.09%) | 2 (7.14%) |  |  |
| School Grade (mean[SD]) | 10.10 [1.23] | 9.82 [1.33] | 10.32 [1.12] | t=1.42 | 0.163 |
| Psychiatric Variables |  |  |  |  |  |
| Depression Severity (KDRS; mean[SD]) | 2.80 [4.41] | 4.41 [5.91] | 1.54 [2.08] | t=-2.18 | **0.039** |
| Mania Severity (KMRS; mean[SD]) | 1.30 [2.31] | 2.09 [3.10] | 0.68 [1.12] | t=-2.03 | 0.052 |
| Mood and Feelings Questionnaire (MFQ) <sup>3</sup> | 8.00 [7.41] | 7.52 [8.07] | 8.27 [6.98] | t=0.33 | 0.740 |
| Child Mania Rating Scale (CMRS) <sup>4</sup> | 1.00 [2.88] | 3.41 [3.10] | 1.52 [2.40] | t=-2.31 | **0.026** |
| Current Psychiatric Diagnoses <sup>5</sup> |  |  |  |  |  |
| Depressive Disorder (%) | 3 (6.00%) | 3 (13.64%) | 0 (0.00%) | Fisher's Exact | 0.079 |
| Anxiety Disorder (%) | 8 (16.00%) | 8 (36.36%) | 0 (0.00%) | Fisher's Exact | **0.001** |
| PTSD (%) | 1 (2.00%) | 1 (4.55%) | 0 (0.00%) | Fisher's Exact | 0.440 |
| ADHD (%) | 5 (10.00%) | 5 (22.73%) | 0 (0.00%) | Fisher's Exact | **0.012** |
| Behavior Disorder (%) | 1 (2.00%) | 1 (4.55%) | 0 (0.00%) | Fisher's Exact | 0.440 |
| Lifetime Psychiatric Diagnoses <sup>5</sup> |  |  |  |  |  |
| Depressive Disorder (%) | 8 (16.00%) | 8 (36.36%) | 0 (0.00%) | Fisher's Exact | **0.001** |
| Anxiety Disorder (%) | 9 (18.00%) | 9 (40.91%) | 0 (0.00%) | Fisher's Exact | **<0.001** |
| PTSD (%) | 1 (2.00%) | 1 (4.55%) | 0 (0.00%) | Fisher's Exact | 0.440 |
| ADHD (%) | 6 (12.00%) | 6 (27.27%) | 0 (0.00%) | Fisher's Exact | **0.005** |
| Behavior Disorder (%) | 1 (2.00%) | 1 (4.55%) | 0 (0.00%) | Fisher's Exact | 0.440 |
| Sleep Variables |  |  |  |  |  |
| Sleep Quality (PSQI-Past Week; mean [SD]) | 3.59 [2.55] | 4.3 [3.08] | 3.04 [1.95] | t=-1.60 | 0.120 |
| Total Actigraphy Days (mean[SD]) | 14.08 [3.97] | 13.64 [3.97] | 14.43 [4.00] | t=0.70 | 0.489 |
| Valid Actigraphy Days (mean[SD]) | 12.16 [4.35] | 12.59 [4.32] | 11.82 [4.43] | t=-0.62 | 0.539 |
| Current Sleep Diagnoses |  |  |  |  |  |
| Insomnia (%) | 7 (14.00%) | 5 (22.73%) | 2 (7.14%) | Fisher's Exact | 0.217 |
| Hypersomnia (%) | 3 (6.00%) | 2 (9.09%) | 1 (3.57%) | *Fisher's Exact* | 0.576 |
| Delayed Sleep Phase Disorder (%) | 1 (2.00%) | 0 (0.00%) | 1 (3.57%) | *Fisher's Exact* | >0.999 |
| Irregular Sleep-Wake Disorder (%) | 7 (14.00%) | 5 (22.70%) | 0 (0.00%) | *Fisher's Exact* | 0.189 |
| Lifetime Sleep Diagnoses |  |  |  |  |  |
| Insomnia (%) | 8 (16.00%) | 6 (27.27%) | 2 (7.14%) | *Fisher's Exact* | 0.063 |
| Hypersomnia (%) | 3 (6.00%) | 2 (9.09%) | 1 (3.57%) | *Fisher's Exact* | 0.571 |
| Delayed Sleep Phase Disorder (%) | 1 (2.00%) | 0 (0.00%) | 1 (3.57%) | *Fisher's Exact* | > 0.999 |
| Irregular Sleep-Wake Disorder (%) | 2 (4.00%) | 2 (9.09%) | 0 (0.00%) | *Fisher's Exact* | 0.181 |

<sup>1</sup> Only 1 participant in the High-risk group was taking psychotropic medication (Nortriptyline).
<sup>2</sup> P-values $\leq 0.05$ are reported in bold characters; P-values between 0.05 and 0.10 are reported in italics.
<sup>3</sup> One participant in the High-Risk group and Two participants in the Low-Risk group were missing the MFQ metric at Baseline.
<sup>4</sup> Three participants in the Low-Risk group were missing the CMRS metric at Baseline.
<sup>5</sup> Depressive Disorder = Persistent Depressive Disorder, Major Depressive Disorder, or Unspecified or Other Specified Depressive Disorder; Anxiety Disorder = Generalized Anxiety Disorder, Obsessive Compulsive Disorder, Panic Disorder, Social Phobia, Specific Phobia, Agoraphobia, Separation Anxiety Disorder; PTSD = Post Traumatic Stress disorder; ADHD = Attention Deficit Hyperactivity Disorder; Behavior Disorder = Oppositional Defiant Disorder, Conduct Disorder.

Low-Risk participants a) had no current or lifetime history of psychiatric conditions based on the Kiddie Schedule for Affective Disorders and Schizophrenia (KSADS(Kaufman et al., 1997)), b) had no first degree family history (parent, sibling) of a psychiatric condition based on the Structured Clinical Interview for the DSM-5, Research Version (SCID(First M.B. et al., 2015)) and Family History Interview (FHI(Andreasen et al., 1977)), and c) were not taking psychotropic or hypnotic medications.

High-Risk participants were a) free of lifetime bipolar disorder, lifetime psychotic disorder, and alcohol/substance use disorders in the past 3 months (KSADS(Kaufman et al., 1997)), b) had at least one parent diagnosed with DSM-5 bipolar disorder, type 1, type 2, or Other Specified (SCID(First M.B. et al., 2015) and/or FHI(Andreasen et al., 1977)), and c) could be taking psychotropic medications if the medication type and dosage were stable for at least 2 months.

### 2.2. Measures

#### 2.2.1. Psychiatric and Sleep Disorders

DSM-5 psychiatric disorders were assessed using the KSADS(Kaufman et al., 1997) in offspring participants, the SCID(First M.B. et al., 2015) in the parent attending the study visit, and the FHI(Andreasen et al., 1977) in first-degrees relatives of the offspring participant. The Pittsburgh Sleep Diagnostic Interview (PSDI) assessed for offspring sleep disorders using DSM-5 criteria.

#### 2.2.2. Depression and Mania severity

The KSADS depression rating scale (KDRS(Axelson et al., 2003)) and Mania rating scale (KMRS(Axelson et al., 2003)) were used to assess symptom severity at Baseline weeks. Depressive and mania symptoms were assessed pre-and post-manipulation using adapted self-report versions of the Mood and Feelings Questionnaire (MFQ(Angold et al., 1995)) and the Child Mania Rating Scale (CMRS(Pavuluri et al., 2006)), referencing symptoms over the past week to enable tracking of short-term symptom changes.

#### 2.2.3. Sleep Diary

Daily self-report sleep diary captured standard data elements recommended by the Consensus Sleep Diary(Carney et al., 2012) (e.g., bedtime, lights off time, sleep latency, wake after sleep onset, number of nocturnal awakenings, sleep offset time, get up time) for each bout of sleep. In addition, participants were asked to classify each sleep bout as a nap or their main sleep interval for the day. Links to the sleep diary were distributed to participants via text message each morning at their preferred time, based on habitual weekday and weekend rise times. Sleep diaries had a 48-hour response window (i.e., participants could only report on their sleep patterns for that day and the prior day).

#### 2.2.4. Actigraphy

A wrist accelerometer (Actiwatch Spectrum; Philips Respironics, Bend, OR) was worn continuously on the non-dominant wrist for 14 days. The Actiwatch Spectrum includes an accelerometer to assess activity and a light sensor composed of color-sensitive photodiodes to measure illuminance of environmental white light (range 0.1-200,000 lux). Light and accelerometer data were collected in 60-sec epochs. Participants were instructed to wear their Actiwatch continuously except in salt water and during activities that may damage the device (e.g., contact sports). For each sleep interval (nap and main), participants were asked to indicate ‘lights off’ (i.e., the beginning of a sleep attempt) and ‘get up’ (i.e., physically getting up from a sleep attempt) via button press on the Actiwatch.

#### 2.2.5. Sleep Estimation

Trained scorers manually identified rest intervals based on a combination of event markers indicated by participants, sleep diary times, and clear changes in activity and environmental light level recorded by the device. Activity-based sleep estimation was performed using a medium wake threshold setting for the main rest interval (10 minutes immobile time for sleep onset and offset). For detection of minor rest intervals (naps), we used a medium wake threshold with a 45-minute minimum(Kanady et al., 2011). Off-wrist and invalid time were automatically converted to Excluded intervals. Actigraphy ‘tracking days’ were defined as noon-to-noon. We implemented additional semi-automated quality assurance procedures using in-house R scripts, including identification of the main rest interval (defined as the longest rest interval each day), removal of invalid sleep intervals containing ≥1 hour of off-wrist time or recording errors(Feliciano et al., 2019; Patel et al., 2015), removal of tracking days with >4 hours non-wear time during active intervals(Patel et al., 2015), time adjustment for daylight savings time, and final visual inspection of sleep intervals on raster plots. The average and standard deviation (variability) of standard sleep variables were extracted: lights off time, sleep latency (minutes between lights off and sleep onset), sleep onset time, minutes awake after sleep onset, sleep offset time, get up time, snooze time (minutes between sleep offset and get up time), time in bed (time between lights off and get up time), total sleep time (time in bed minus sleep latency, wake after sleep onset, and snooze time). Midsleep was calculated as the midpoint between sleep onset and offset. Primary measures were sleep duration (total sleep time) and midsleep variability; lights off time and get up time were used in exploratory analyses.

#### 2.2.6. Rest-activity rhythms (RARs)

Non-parametric RAR outcomes were derived with the nParACT R package (Smagula et al., 2018). Non-wear segments were automatically imputed using the median time of a day(Weed et al., 2022), which relies on data from non-missing days at the corresponding timepoints of missing data and is recommended for periods of missing data shorter than 24 hours(Weed et al., 2022). We calculated three primary outcomes: 1) Intradaily Variability (IV), which reflects how much activity changes from hour to hour relative to overall activity variability; higher values indicate more fragmented RAR; 2) Interdaily Stability (IS), which reflects consistency of activity patterns from day-to-day; higher values indicate greater RAR stability; and 3) Relative Amplitude (RA), which reflects the average activity ratio of the active 10 hours relative to the least active 5 hours of the day; higher values indicate greater RAR amplitude. Additionally, as a secondary outcome, we calculated the Circadian Function Index (CFI)(Ortiz-Tudela et al.), a composite measure of circadian activity rhythm robustness; it is calculated as the average of IS, RA, and a transformed IV component (1−IV/2). The CFI can range from 0 (absence of circadian rhythmicity) and 1 (a robust circadian rhythm).

### 2.3. Procedures

#### 2.3.1 Clinical Evaluation

Participants were recruited via community advertisement, local clinics, and a participant registry. Adolescent participants (and a parent, if <18yr) completed consent/assent and an evaluation to determine study eligibility that included psychiatric diagnostic interviews (SCID, KSADS, FHI), a sleep diagnostic interview, and questionnaires.

#### 2.3.2 Sleep Monitoring

Eligible participants were invited to complete 2 weeks of continuous actigraphy monitoring and an online sleep diary each morning. Sleep monitoring occurred during the school year only. At the end of the sleep monitoring period, self-rated depressive and mania symptoms were collected using the adapted versions of the MFQ and CMRS respectively.

A subset of high-risk participants with >2 hours difference in bedtime and/or risetime between the weekday and weekend were invited to participate in an additional two-week sleep stabilization manipulation. Actigraphy monitoring and online sleep diary were also collected during the stabilization manipulation weeks. At the end of the two-week sleep stabilization manipulation, self-rated depressive and mania symptoms were collected using the adapted versions of the MFQ and CMRS respectively, as clinician-administered assessments were not available at the post-manipulation time point.

#### 2.3.3 Sleep Stabilization Manipulation

Sleep stabilization procedures were distilled from behavioral sleep interventions developed for inter-episode BD (CBTI-BP(Harvey et al., 2015)) and teens (Transdiagnostic Sleep and Circadian Intervention - TranS-C(Harvey, In Press)). CBTI-BP was determined to be safe, tolerable, and efficacious in inter-episode BD(Harvey et al., 2015; Kaplan and Harvey, 2013). TranS-C provides adaptations for treating sleep problems in the context of adolescence(Harvey, In Press). Both packages draw from evidence-based therapies that target variable sleep patterns in psychiatric samples, including CBT for insomnia and social rhythm therapy (e.g.,(Goldstein et al., 2014b; Wu et al., 2015)). Stability of sleep timing *and* duration was targeted in this pilot study(Manber et al., 1996), as these constructs anchor sleep schedules.

An individualized behavioral plan was devised with participants and their caregivers that included, when appropriate: a) a prescribed sleep schedule (bedtime, risetime, 9hr sleep opportunity); b) modified stimulus control recommendations (no naps, limit activities in bed to sleep); c) a nighttime wind-down routine (relaxing/sleep-enhancing activities in low-light); d) a morning wake-up routine (behavioral plan to address sleep inertia). These procedures facilitated regular sleep timing and opportunity.

Adherence to the protocol was reinforced in several ways. First, motivational interviewing facilitated setting a realistic sleep schedule and discussion of anticipated barriers to the protocol. Second, a phone check-in was made after the first week to address any adherence barriers(Buysse et al., 2011) and assess clinical status (self-rated sleepiness & clinician-rated mania and depressive symptoms). Third, the online sleep diary was monitored daily by study staff. Lastly, at lab visit 3, bonus compensation was provided for sleep schedule adherence based on sleep diary and light/motion data from actigraphy.

### 2.4. Statistical approach

Baseline sleep/RAR actigraphy measures were calculated across the two weeks prior to the sleep stabilization manipulation. Post-manipulation metrics were evaluated separately for manipulation week 1 and manipulation week 2, as we hypothesized that adherence to the sleep manipulation might wane after the first week without a full therapeutic intervention to support sustained behavior change. In addition, given that the distribution of depression (KDRS) and mania (KMRS) severity across participants at Baseline, as well as self-rated depressive symptoms pre-and post-manipulation (MFQ), were non-normal, these scores were log transformed before statistical analyses. In contrast, self-rated mania scores (CMRS) met the assumption of normality, as indicated by the Shapiro-Wilk test. Age, sex at birth, and actigraphy variables (total tracking days, ratio of weekdays to weekend days) were included as covariates in all models. False Discovery Rate (FDR(Benjamini and Hochberg, 1995)) was used to account for multiple comparisons.

#### 2.4.1 Baseline comparisons

Analysis of Covariance (ANCOVA) models investigated between-group differences (High-Risk versus Low-Risk) in sleep duration variability, midsleep variability, and CFI at Baseline.

#### 2.4.2. Moderation effects

ANCOVA models examined whether group (High-Risk versus Low-Risk) moderated the effect of independent variables (sleep duration variability, midsleep variability, or CFI) on the severity of symptoms (depression – KDRS; mania – KMRS) at Baseline. Moderation effects were tested through an interaction term between group and independent variables in the models. Models were performed independently. Significant interactions were further explored by comparing estimated marginal means(Lenth, 2020).

#### 2.4.3. Sleep stabilization effects

Linear mixed effects models investigated the effect of week (Baseline, Manipulation week 1, and Manipulation week 2) on our outcomes of interest (sleep duration variability, midsleep variability, and CFI). Additionally, linear mixed effects models investigated the effect of manipulation (pre-*versus* post-manipulation) on self-rated depressive (MFQ) and mania (CMRS) symptoms.

#### 2.4.4. Exploratory analyses

Models parallel to those described in 2.4.1 and 2.4.3 explored Baseline differences and sleep manipulation effects on other sleep patterns and RAR metrics: sleep duration average, midsleep average, lights off time and get up time. Additional models investigated whether sleep duration average and midsleep average had a moderation effect in models parallel to those described in 2.4.2.

Finally, linear regression models were used to examine the association between changes (Δ Baseline *minus* Manipulation) in sleep/RAR measures (sleep duration variability, midsleep variability, and CFI) and self-rated symptoms (CMRS and MFQ). Since the clinical assessments were collected at the end of the 2-week sleep stabilization manipulation, changes in sleep/RAR were calculated as the difference between average values during the Baseline weeks and the manipulation weeks.

## 3. RESULTS

### 3.1. Sample characteristics

Adolescents in the High-Risk group showed higher depression severity (KDRS) and reported more depression (lifetime), anxiety (lifetime and current), and ADHD (lifetime) than adolescents in the Low-Risk group. They also reported more mania symptoms (CMRS) than the Low-Risk group. There were no other differences between groups (**Table 1**). Eleven participants from the High-Risk group were invited to complete a 2-week sleep stabilization manipulation. There were no differences between the original High-Risk sample (N=22) and the subsample invited for sleep stabilization (**Supplemental Table 1**).

### 3.2. Baseline comparisons

Baseline sleep duration variability, midsleep variability, and CFI did not significantly differ between the High-Risk versus Low-Risk groups. Additionally, there were no Baseline differences in the other sleep and RAR metrics (**Table 2**).

**Table 2.** Between-group differences in sleep and RAR metrics at Baseline.

| **Sleep and RAR metrics included in the main models** |  |  |  |  |  |  |
| --- | --- | --- | --- | --- | --- | --- |
| Variable | High-Risk | Low-Risk | F | P <sup>1</sup> | Q <sup>1,2</sup> | Cohen's d <sup>3</sup> |
| Sleep duration - variability | 94.81 [49.43] | 81.72 [47.73] | 0.86 | 0.359 | 0.359 | 0.28 |
| Midsleep - variability | 99.75 [71.88] | 73.52 [39.04] | 2.59 | 0.115 | 0.201 | 0.49 |
| CFI | 0.63 [0.09] | 0.67 [0.07] | 2.33 | 0.134 | 0.201 | 0.46 |
| **Other sleep and RAR metrics included in exploratory analyses** |  |  |  |  |  |  |
| Variable | High-Risk | Low-Risk | F | P <sup>1</sup> | Q <sup>1,2</sup> | Cohen's d <sup>3</sup> |
| Sleep duration - average | 402.53 [41.06] | 391.89 [41.67] | 0.81 | 0.373 | 0.579 | 0.27 |
| Midsleep - average | 238.01 [74.8] | 226.6 [75.97] | 0.31 | 0.579 | 0.579 | 0.17 |
| Lights off time - average | 1412.11 [71.94] | 1427.9 [82.43] | 0.57 | 0.452 | 0.579 | 0.23 |
| Get up time - average | 484.77 [74.81] | 466.2 [81.31] | 0.77 | 0.384 | 0.579 | 0.27 |

Abbreviations: RAR - rest-activity rhythms; CFI - Circadian function index.
<sup>1</sup> P-values $\leq 0.05$ are reported in bold characters; P-values between 0.05 and 0.10 are reported in italics.
<sup>2</sup> Q-values indicate P-values after False Discovery Rate (FDR) correction for multiple comparisons.
<sup>3</sup> Cohen's d benchmarks: small = 0.2, medium = 0.5, large = 0.8.

### 3.3. Associations with mood

The association between sleep duration variability and depression severity (KDRS) was moderated by group status (**Table 3**). Specifically, in the High-Risk group, greater sleep duration variability was associated with higher depression severity (**Figure 1 – Panel A**). This association was not observed in the Low-Risk group.

**Figure 1.**
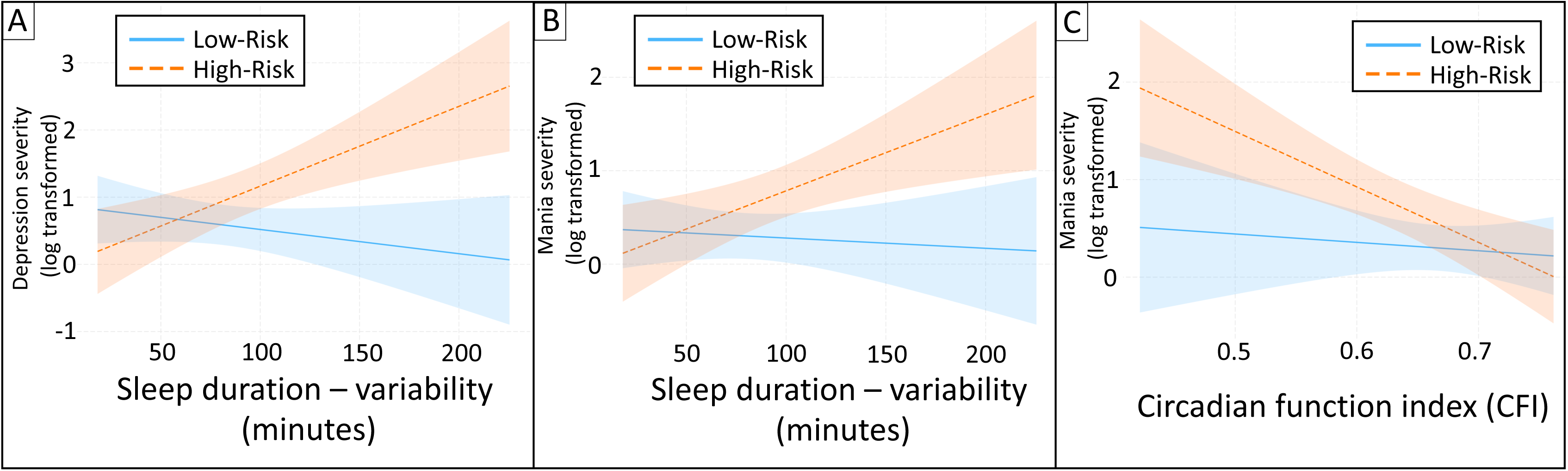
Moderation effect of group (High-Risk versus Low-Risk) in the effects of sleep and RAR instability on symptom severity. Figure 1 shows scatter plots with the relationship between sleep duration variability and depression severity (KDRS; Panel A), sleep duration variability and mania severity (KMRS; Panel B), and CFI and mania severity (KMRS; Panel C). The x-axis shows the sleep variables, and the y-axis shows the log-transformed scores of symptom severity (KDRS for depression and KMRS for mania). These scores were log transformed before statistical analyses due to non-normal distribution. The relationships in the High-Risk group are demonstrated in dotted regression lines while the relationships in Low-Risk groups are demonstrated in solid regression lines. The light-colored area around the regression line represents the 95% CI. Only the relationships in the High-Risk group were statistically significant (P<0.05). Abbreviations: RAR – rest-activity rhythms; CFI – Circadian Function Index; KDRS - KSADS depression rating scale; KMRS – KSADS Mania rating scale.

**Table 3.** Between-group differences regarding the impact of sleep and RAR instability on depression and mania severity (moderation effects).

| Moderation effect results |  |  |  |  |  |  |  |
| --- | --- | --- | --- | --- | --- | --- | --- |
| Outcome | Independent variable | F | $\eta^2$ <sup>1</sup> | P <sup>2</sup> | Q <sup>2,3</sup> | | |
| Depression Severity<br>(KDRS) <sup>4</sup> | Sleep duration - variability | 20.67 | 0.330 | **<0.001** | **<0.001** |  |  |
|  | Midsleep - variability | 2.16 | 0.049 | 0.149 | 0.163 |  |  |
|  | Circadian function index | 2.02 | 0.046 | 0.163 | 0.163 |  |  |
| Mania Severity<br>(KMRS) <sup>4</sup> | Sleep duration - variability | 5.71 | 0.120 | **0.021** | **0.044** |  |  |
|  | Midsleep - variability | 2.14 | 0.048 | 0.151 | 0.163 |  |  |
|  | Circadian function index (CFI) | 5.65 | 0.119 | **0.022** | **0.044** |  |  |
| Estimated marginal means for significant moderation effects |  |  |  |  |  |  |  |
| Outcome | Independent variable | Group <sup>1</sup> | $\beta$ <sup>1</sup> | SE <sup>1</sup> | DF <sup>1</sup> | LLCI <sup>1</sup> | ULCI <sup>1</sup> |
| Depression Severity<br>(KDRS) <sup>4</sup> | Sleep duration –<br>variability | Low-Risk | -0.006 | 0.003 | 42 | -0.012 | -0.001 |
|  |  | **High-Risk** | **0.012** | **0.003** | **42** | **0.006** | **0.018** |
| Mania Severity<br>(KMRS) <sup>4</sup> | Sleep duration -<br>variability | Low-Risk | -0.002 | 0.004 | 42 | -0.009 | 0.005 |
|  |  | **High-Risk** | **0.011** | **0.004** | **42** | **0.003** | **0.019** |
|  | Circadian function<br>index (CFI) | Low-Risk | -0.726 | 2.331 | 42 | -5.430 | 3.977 |
|  |  | **High-Risk** | **-8.236** | **2.125** | **42** | **-12.525** | **-3.940** |

Abbreviations: RAR - rest-activity rhythms; KDRS - KSADS depression rating scale; KMRS – KSADS Mania rating scale; SE – Standard error; DF – degrees of freedom; LLCI – Lower limit confidence interval; ULCI – Upper limit confidence interval.
<sup>1</sup> $\eta^2$ effect size benchmarks: negligible ( $\eta^2 \geq 0.01$ ), small ( $0.01 \leq \eta^2 < 0.06$ ), medium ( $0.06 \leq \eta^2 < 0.14$ ), and large ( $\eta^2 \geq 0.14$ ) partial effect sizes.
<sup>2</sup> P-values $\leq 0.05$ are reported in bold characters; P-values between 0.05 and 0.10 are reported in italics.
<sup>3</sup> Q-values indicate P-values after False Discovery Rate (FDR) correction for multiple comparisons.
<sup>4</sup> The distribution of depression (KDRS) and mania (KMRS) severity across participants was not normally distributed; these scores were log transformed before statistical analyses.
<sup>5</sup> Associations with confidence intervals that do not include 0 are statistically significant and are reported in bold characters.

Similarly, associations between sleep duration variability and CFI with mania severity (KMRS) also differed significantly between groups (**Table 3**). In the High-Risk group, greater sleep duration variability and lower CFI were associated with higher mania severity **(Figure 1 – Panels B-C**). These associations were not observed in the Low-Risk group.

In exploratory analyses, the association between midsleep and depression severity (KDRS) was moderated by group status. We found that later midsleep was associated with higher depression severity in High-Risk adolescents. Additional exploratory analyses indicated that the association between sleep duration and midsleep with mania severity (KMRS) was also moderated by group status. In the High-Risk group, shorter sleep duration and later midsleep were associated with higher mania severity. These relationships were not present in Low-Risk adolescents (**Supplemental Table 2; Supplemental Figure 1**).

### 3.4. Sleep stabilization effects

The sleep stabilization manipulation significantly decreased sleep duration variability and midsleep variability, and increased CFI (**Table 4**; **Figure 2**). Pairwise comparisons revealed a significant effect during manipulation week 1 (relative to Baseline) across all metrics. However, by manipulation week 2, stabilization effects waned, with only midsleep variability remaining significantly lower than Baseline values. Notably, pairwise comparisons between manipulation weeks 1 and 2 showed no significant differences, suggesting that some of the initial changes persisted over time (**Supplemental Table 3**).

**Figure 2.**
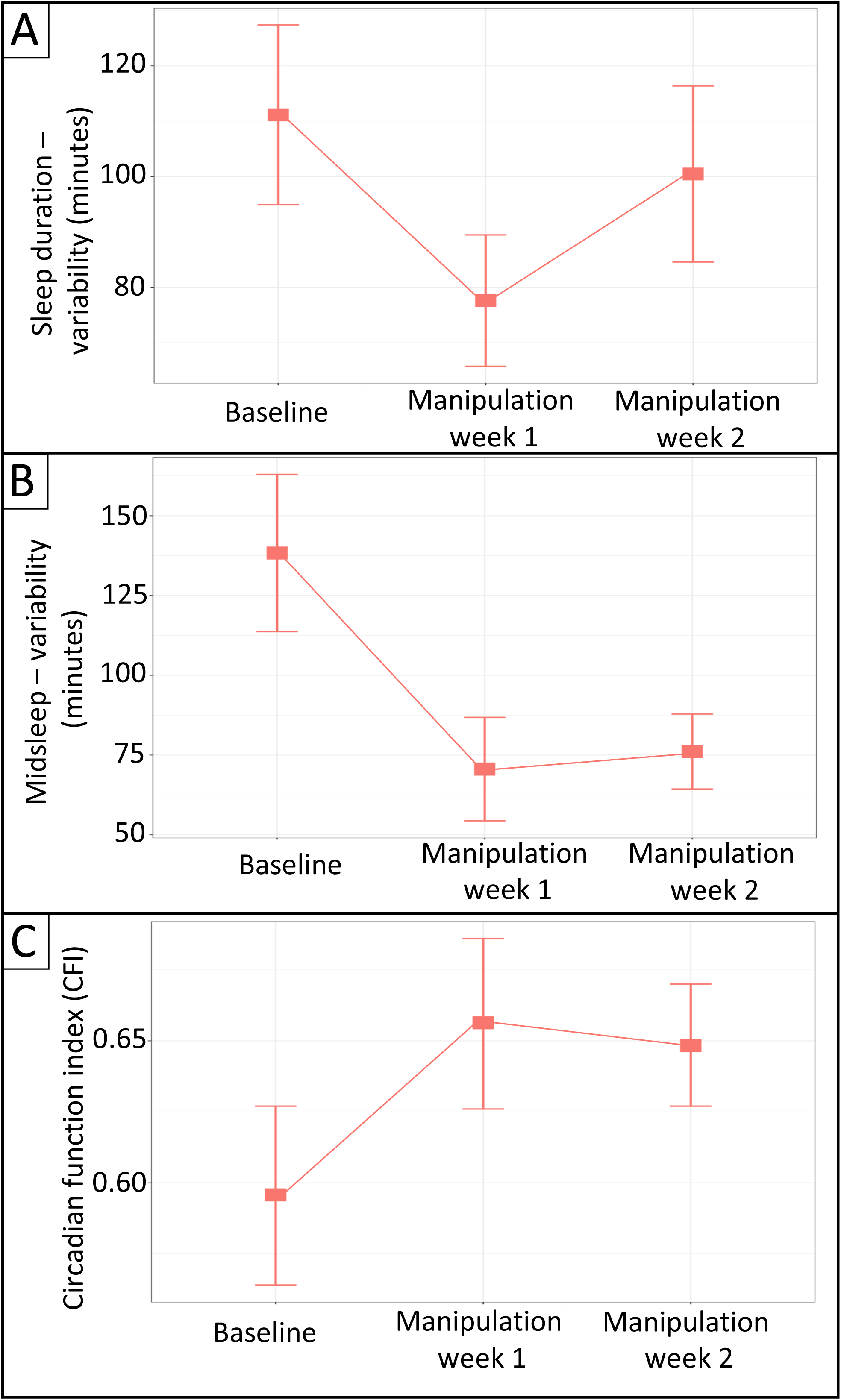
Effect of sleep stabilization manipulation on sleep and RAR instability. Figure 2 shows the differences in sleep duration variability (Panel A), midsleep variability (Panel B), and CFI (Panel C) across time (Baseline, Manipulation week 1, and Manipulation week 2) in 11 High-risk adolescents who took part in the stabilization manipulation protocol. X-axis shows the timepoints and the y-axis shows the sleep and RAR metrics. Error bars represent the standard error. Abbreviations: RAR – rest-activity rhythms; CFI – Circadian Function Index

**Table 4.** Association between sleep stabilization manipulation, sleep/RAR instability, and mood symptoms (MFQ and CMRS).

| Sleep stabilization manipulation and sleep/RAR instability |  |  |  |  |  |  |  |
| --- | --- | --- | --- | --- | --- | --- | --- |
| Outcome | Baseline weeks Mean [SD] | Manipulation week 1 Mean [SD] | Manipulation week 2 Mean [SD] | F | $\eta^2$<sup>1</sup> | P<sup>2</sup> | Q<sup>2,3</sup> |
| Sleep duration - variability (minutes) | 111.15 [53.83] | 77.6 [39.35] | 100.47 [52.72] | 12.93 | 0.590 | **<0.001** | **<0.001** |
| Midsleep - Variability (minutes) | 138.32 [81.69] | 70.59 [53.76] | 76.1 [39] | 7.23 | 0.446 | **0.005** | **0.008** |
| Circadian function index | 0.60 [0.11] | 0.66 [0.10] | 0.65 [0.07] | 3.72 | 0.293 | **0.044** | **0.044** |
| Sleep stabilization manipulation and mood symptoms |  |  |  |  |  |  |  |
| Outcome | Pre-manipulation | Post-manipulation | F | $\eta^2$<sup>1</sup> | P<sup>2</sup> | Q<sup>2,3</sup> | |
| Mood and Feelings Questionnaire (MFQ)<sup>4</sup> | 8.00 [7.40] | 9.09 [7.02] | 0.08 | 0.010 | 0.780 | 0.780 |  |
| Child Mania Rating Scale (CMRS)<sup>5</sup> | 4.55 [3.59] | 3.20 [3.08] | 6.17 | 0.469 | **0.042** | *0.080* |  |

Abbreviations: RAR - rest-activity rhythms.
<sup>1</sup> $\eta^2$ effect size benchmarks: negligible ( $\eta^2 \geq 0.01$ ), small ( $0.01 \leq \eta^2 < 0.06$ ), medium ( $0.06 \leq \eta^2 < 0.14$ ), and large ( $\eta^2 \geq 0.14$ ) partial effect sizes.
<sup>2</sup> P-values $\leq 0.05$ are reported in bold characters; P-values between 0.05 and 0.10 are reported in italics.
<sup>3</sup> Q-values indicate P-values after False Discovery Rate (FDR) correction for multiple comparisons.
<sup>4</sup> The distribution of the MFQ across participants was not normally distributed; these scores were log transformed before statistical analyses.
<sup>5</sup> One participant was missing CMRS data in the Post-manipulation self-report.

In exploratory analyses of the effects of sleep stabilization manipulation on other sleep/RAR metrics (sleep duration average, midsleep average, lights off time and get up time), only get up time showed a significant effect, but that did not survive multiple correction comparisons (**Supplemental Table 4**).

Finally, we observed a reduction in self-rated mania symptoms (CMRS) following the sleep stabilization manipulation, although this effect did not survive correction for multiple comparisons (**Table 4**). Effects of the manipulation on self-rated depression (MFQ) were not significant. Additional exploratory models revealed that pre-to post-manipulation improvement in circadian function (CFI) was associated with a decrease in self-rated depressive symptoms; an increase sleep duration was associated with a reduction in self-rated mania symptoms; and later get-up time was associated with a reduction in self-rated mania symptoms (**Supplemental Table 5**). However, these associations did not survive correction for multiple comparisons.

## 4. DISCUSSION

We examined whether High-Risk adolescents are more vulnerable to sleep/RAR instability than their Low-Risk peers and whether a brief, at-home sleep manipulation protocol stabilized sleep/RAR and mood symptoms. Supporting our first hypothesis, greater sleep and RAR instability was associated with greater depressive and mania symptoms in High-Risk adolescents—a relationship not observed in the Low-Risk group. To the best of our knowledge, this is the first study to demonstrate that sleep and RAR instability are differentially linked to mood symptoms based on familial risk for BD during adolescence. Additionally, supporting our second hypothesis, the stabilization manipulation protocol effectively reduced sleep/RAR instability and self-rated mania symptoms in the High-Risk group, although the latter did not survive multiple comparison correction.

Despite finding no between-group differences (Low-Risk *versus* High-Risk) in sleep/RAR instability, moderation analyses revealed significantly stronger links with mood symptoms in High-Risk adolescents than in their Low-Risk peers. While previous studies in community samples have linked sleep instability to greater symptom severity, few have specifically examined adolescents with a familial risk for BD(Bei et al., 2017; Mathew et al., 2023). Within High-Risk adolescents, the two facets of sleep/RAR instability, sleep duration variability and CFI, showed partially overlapping but distinct links to mood severity. Greater sleep duration variability predicted higher severity of both depressive and mania symptoms. In contrast, poorer circadian function (lower CFI), which integrates interdaily stability, relative amplitude, and intradaily variability(Ortiz-Tudela et al.), was selectively associated with greater mania severity. This pattern suggests that, while sleep instability acts as a general amplifier of mood dysregulation, specific circadian disruptions may lie closer to the mania end of the spectrum, offering a more targeted early-warning signal. Altogether, our findings suggest that sleep/RAR instability functions less as group-level biomarker and more as individual-level amplifier of existing vulnerability—compounding the vulnerability already present during adolescence(Carskadon, 2011; Cole et al., 1992). In this context, High-Risk adolescents face a double burden: developmental susceptibility to sleep and mood instability coupled with inherited biological vulnerabilities related to BD, potentially accelerating trajectories toward disorder onset. Importantly, even *normative* shifts in sleep/RAR instability may confer risk for adolescents with familial risk for BD. These findings position sleep/RAR instability as an early marker of behavioral vulnerabilities and potential intervention target. However, future longitudinal studies are needed to evaluate the impact of sleep/RAR instability on mood trajectories. Finally, these findings highlight the need to better understand sources of variability in adolescent sleep/RAR stability, particularly in those at familial risk for BD. Contextual factors such as inconsistent daily routines, academic pressure, and social stressors(Carskadon, 2011; Cole et al., 1992), while common in adolescence, may have a disproportionately destabilizing effect among High-Risk youth, underscoring the importance of attending to these modifiable influences in prevention efforts.

Importantly, our sleep manipulation protocol helped stabilize the key components of sleep patterns and RAR that were linked to mood symptoms in High-Risk adolescents, bridging the gap between identifying biological vulnerabilities and testing feasible, non-invasive interventions. Grounded in evidence-based approaches(Goldstein et al., 2014a; Harvey et al., 2015), this protocol offers a novel way to move beyond correlational studies in high-risk samples and probe the mechanisms linking sleep instability to BD risk. Following the sleep stabilization manipulation, we observed a reduction in self-rated mania symptoms (CMRS) suggesting potential benefits of targeting sleep/RAR even for early subthreshold mania features. Although exploratory, additional models also revealed that improvements in circadian function (CFI) were associated with decreases in depressive symptoms, while increases in sleep duration and later get-up times were each associated with reductions in self-rated mania symptoms. Of note, these findings link CFI to depression, suggesting that CFI may exert effects on other mood symptoms. However, these associations involving self-reported mood symptoms did not survive correction for multiple comparisons and, given our small pilot sample, should be interpreted with caution. They may reflect differences in sensitivity or scope between self-reports and clinician ratings, with each capturing distinct facets of mood symptomatology. Future research is still needed to clarify the neural mechanisms that contribute to the link between sleep/RAR instability and mood in High-Risk adolescents, but our findings offer promising evidence that brief, home-based stabilization protocols are one avenue to probe mechanistic brain-behavior associations.

Despite the strengths of this study, some limitations should be noted. First, although our sample was well-characterized in terms of familial risk, the generalizability of findings may be constrained by the small sample size, and demographic or regional factors. Future studies with larger, more diverse samples and additional physiological or neuroimaging data could help clarify the broader applicability and underlying mechanisms of these effects. Second, because our Low-Risk group was comprised of youth without parental history of psychiatric conditions, we cannot rule out that adolescents *any* familial psychiatric history may be more vulnerable to effects of poor sleep on mental health. Third, mood symptoms following sleep stabilization were assessed using self-report measures rather than clinician-rated assessments, which may be influenced by reporting biases. Future studies should incorporate clinician-rated assessments at multiple time points. Finally, while the results suggest promising effects of the manipulation in High-Risk adolescents, we did not include a control arm involving Low-Risk adolescents or alternative manipulation conditions (e.g., sleep extension, sleep monitoring-only, etc.). This limits our ability to determine whether the observed improvements were due to the specific effects of the stabilization protocol or to other factors.

In sum, this study provides novel evidence that adolescents at familial risk for BD show stronger associations between sleep/RAR instability and mood symptoms than typically developing peers, and that a brief, home-based sleep manipulation could be applied to better understand mechanistic relationships between sleep/RAR instability. By linking sleep and circadian dysregulation to mood symptom expression—and demonstrating that these patterns can be modified through feasible, non-invasive interventions—our findings point to sleep and RAR as both a risk marker and a modifiable target for early intervention. Our results lend further support the use of preventative or early intervention strategies for BD that target sleep/RAR instability in High-Risk adolescents and support a methodological shift from correlational to mechanistic studies of sleep-based risk processes in high-risk cohorts.

## Data Availability

Data may be made available from the corresponding author upon reasonable request with the permission of the principal investigators and ethics board approval.

## ACKNOWLEDGEMENTS

This work was supported by the National Institute of Mental Health (K01MH111953; PI: Soehner; R01MH124828; PI: Soehner). This funding agency was not involved in the design, analysis, and interpretation of the data, or the preparation, review, or approval of the manuscript.

## AUTHOR CONTRIBUTIONS

João Paulo Lima Santos: Conceptualization, Formal analysis, and Writing - Original Draft. Lauren Keller: Writing - Review & Editing. Amy G. Hartman: Writing - Review & Editing. Simey Chan: Data Curation and Writing - Review & Editing. Mary L. Phillips: Writing - Review & Editing. Adriane M. Soehner: Conceptualization, Formal analysis, Writing - Review & Editing, Supervision, Project administration, and Funding acquisition.

