## Supplement for "Unstable sleep and rest-activity rhythms in adolescents at-risk for bipolar disorder: links to mood symptoms and the effect of sleep stabilization"

### SUPPLEMENTAL MATERIALS

#### SUPPLEMENTAL TABLES

**Supplemental Table 1. Demographic, clinical, and sleep characteristics in the High-Risk sample invited for sleep stabilization manipulation (N=11).**

| *Variable* | **High-Risk (N=22)** | **High-Risk invited for sleep stabilization (N=11)** | **Between-group differences Statistics [1]** | |
|---|---|---|---|---|
| **Demographics** | | | | |
| Age (Yr; mean[SD]) | 15.87 [1.33] | 15.88 [1.25] | t=0.01 | 0.992 |
| Sex at birth (% Female) | 15 (68.18%) | 8 (72.73%) | $x^2$<0.01 | >0.999 |
| Ethnicity (% Non-Hispanic) | 21 (95.45%) | 11 (100.00%) | $x^2$<0.01 | >0.999 |
| Race | | | | |
| White (%) | 15 (68.18%) | 5 (45.45%) | | |
| Asian (%) | 0 (0.00%) | 0 (0.00%) | $x^2$=5.47 | *0.065* |
| Black (%) | 5 (22.73%) | 4 (36.36%) | | |
| Multiple (%) | 2 (9.09%) | 2 (18.18%) | | |
| School Grade (mean[SD]) | 9.82 [1.33] | 9.91 [1.22] | t=0.31 | 0.758 |
| **Psychiatric Variables** | | | | |
| Depression Severity (KDRS; mean[SD]) | 4.41 [5.91] | 6.09 [6.12] | t=1.36 | 0.189 |
| Mania Severity (KMRS; mean[SD]) | 2.09 [3.10] | 3.18 [3.74] | t=1.73 | 0.105 |
| Current Psychiatric Diagnoses [2] | | | | |
| Depressive Disorder (%) | 3 (13.64%) | 3 (27.27%) | *Fisher's Exact* | 0.214 |
| Anxiety Disorder (%) | 8 (36.36%) | 6 (54.55%) | *Fisher's Exact* | 0.183 |
| PTSD (%) | 1 (4.55%) | 1 (9.09%) | *Fisher's Exact* | >0.999 |
| ADHD (%) | 5 (22.73%) | 4 (36.36%) | *Fisher's Exact* | 0.311 |
| Behavior Disorder (%) | 1 (4.55%) | 1 (9.09%) | *Fisher's Exact* | >0.999 |
| Lifetime Psychiatric Diagnoses [2] | | | | |
| Depressive Disorder (%) | 8 (36.36%) | 6 (54.55%) | *Fisher's Exact* | 0.183 |
| Anxiety Disorder (%) | 9 (40.91%) | 6 (54.55%) | *Fisher's Exact* | 0.387 |
| PTSD (%) | 1 (4.55%) | 1 (9.09%) | *Fisher's Exact* | >0.999 |
| ADHD (%) | 6 (27.27%) | 4 (36.36%) | *Fisher's Exact* | 0.635 |
| Behavior Disorder (%) | 1 (4.55%) | 1 (9.09%) | *Fisher's Exact* | >0.999 |
| **Sleep Variables** | | | | |
| Sleep Quality (PSQI-Past Week; mean [SD]) | 4.3 [3.08] | 5.27 [2.97] | t=-1.60 | 0.121 |
| Total Actigraphy Days (mean[SD]) | 13.64 [3.97] | 13.82 [3.84] | t=0.70 | 0.836 |
| Valid Actigraphy Days (mean[SD]) | 12.59 [4.32] | 12.64 [4.39] | t=-0.62 | 0.962 |
| Current Sleep Diagnoses | | | | |

| | | | | |
|---|---|---|---|---|
| Insomnia (%) | 5 (22.73%) | 4 (36.36%) | *Fisher's Exact* | 0.311 |
| Hypersomnia (%) | 2 (9.09%) | 1 (9.09%) | *Fisher's Exact* | >0.999 |
| Delayed Sleep Phase Disorder (%) | 0 (0.00%) | 0 (0.00%) | *Fisher's Exact* | >0.999 |
| Irregular Sleep-Wake Disorder (%) | 5 (22.70%) | 2 (18.18%) | *Fisher's Exact* | 0.476 |
| Lifetime Sleep Diagnoses | | | | |
| Insomnia (%) | 6 (27.27%) | 5 (45.45%) | *Fisher's Exact* | 0.149 |
| Hypersomnia (%) | 2 (9.09%) | 1 (9.09%) | *Fisher's Exact* | >0.999 |
| Delayed Sleep Phase Disorder (%) | 0 (0.00%) | 0 (0.00%) | *Fisher's Exact* | >0.999 |
| Irregular Sleep-Wake Disorder (%) | 2 (9.09%) | 2 (18.18%) | *Fisher's Exact* | 0.476 |

[1] P-values $\leq 0.05$ are reported in bold characters; P-values between 0.05 and 0.10 are reported in italics.

[2] Depressive Disorder = Persistent Depressive Disorder, Major Depressive Disorder, or Unspecified or Other Specified Depressive Disorder; Anxiety Disorder = Generalized Anxiety Disorder, Obsessive Compulsive Disorder, Panic Disorder, Social Phobia, Specific Phobia, Agoraphobia, Separation Anxiety Disorder; PTSD = Post Traumatic Stress disorder; ADHD = Attention Deficit Hyperactivity Disorder; Behavior Disorder = Oppositional Defiant Disorder, Conduct Disorder.

**Supplemental Table 2. Between-group differences regarding the impact of other sleep and RAR metrics on depression and mania severity (moderation effects). Between-group differences regarding the impact of other sleep and RAR metrics on depression and mania severity (moderation effects).**

**Moderation effect results**

| Outcome | Independent variable | F | η2 [1] | P [2] | Q [2,3] |
|---|---|---|---|---|---|
| Depression Severity (KDRS)[4] | Sleep duration (minutes) - average | 0.38 | 0.009 | 0.539 | 0.539 |
| | Midsleep (minutes) - average | 10.33 | 0.197 | **0.003** | **0.012** |
| Mania Severity (KMRS)[4] | Sleep duration (minutes) - average | 5.43 | 0.115 | **0.025** | **0.033** |
| | Midsleep (minutes) - average | 6.33 | 0.131 | **0.016** | **0.032** |

**Estimated marginal means for significant moderation effects**

| Outcome | Independent variable | Group [1] | β [1] | SE [1] | DF [1] | LLCI [1] | ULCI [1] |
|---|---|---|---|---|---|---|---|
| Depression Severity (KDRS) [4] | Midsleep - average | Low-Risk | -0.003 | 0.002 | 42 | -0.007 | 0.001 |
| | | **High-Risk** | **0.006** | **0.002** | **42** | **0.002** | **0.011** |
| Mania Severity (KMRS) [4] | Sleep duration (minutes) - average | Low-Risk | 0.003 | 0.004 | 42 | -0.006 | 0.011 |
| | | **High-Risk** | **-0.013** | **0.005** | **42** | **-0.023** | **-0.003** |
| | Midsleep (minutes) - average | Low-Risk | -0.003 | 0.002 | 42 | -0.008 | 0.002 |
| | | **High-Risk** | **0.236** | **0.003** | **42** | **0.001** | **0.012** |

Abbreviations: RAR - rest-activity rhythms; KDRS - KSADS depression rating scale; KMRS – KSADS Mania rating scale; SE – Standard error; DF – degrees of freedom; LLCI – Lower limit confidence interval; ULCI – Upper limit confidence interval.

[1] η2 effect size benchmarks: negligible ($\eta2 \geq 0.01$), small ($0.01 \leq \eta2 < 0.06$), medium ($0.06 \leq \eta2 < 0.14$), and large ($\eta2 \geq 0.14$) partial effect sizes.

[2] P-values ≤ 0.05 are reported in bold characters; P-values between 0.05 and 0.10 are reported in italics.

[3] Q-values indicate P-values after False Discovery Rate (FDR) correction for multiple comparisons.

[4] The distribution of depression (KDRS) and mania (KMRS) severity across participants was not normally distributed; these scores were log transformed before statistical analyses.

[5] Associations with confidence intervals that do not include 0 are statistically significant and are reported in bold characters.

**Supplemental Table 3. Pairwise comparisons between Baseline, manipulation week 1, and manipulation week 2.**

| **Pairwise comparisons - Baseline versus Manipulation week 1** | | | |
|---|---|---|---|
| Outcome | F | η2 [1] | P [2] |
| Sleep duration (minutes) - variability | 11.73 | 0.594 | **0.009** |
| Midsleep (minutes) - variability | 13.23 | 0.623 | **0.007** |
| Circadian function index | 6.12 | 0.433 | **0.038** |
| **Pairwise comparisons - Baseline versus Manipulation week 2** | | | |
| Outcome | F | η2 [1] | P [2] |
| Sleep duration (minutes) - variability | 0.5 | 0.059 | 0.500 |
| Midsleep (minutes) - variability | 7.58 | 0.487 | **0.025** |
| Circadian function index | 5.06 | 0.387 | *0.055* |
| **Pairwise comparisons - Manipulation week 1 versus Manipulation week 2** | | | |
| Outcome | F | η2 [1] | P [2] |
| Sleep duration (minutes) - variability | 3.87 | 0.326 | *0.085* |
| Midsleep (minutes) - variability | 0.27 | 0.033 | 0.616 |
| Circadian function index | 0.26 | 0.030 | 0.627 |

[1] η2 effect size benchmarks: negligible ($\eta2 \geq 0.01$), small ($0.01 \leq \eta2 < 0.06$), medium ($0.06 \leq \eta2 < 0.14$), and large ($\eta2 \geq 0.14$) partial effect sizes.

[2] P-values ≤ 0.05 are reported in bold characters; P-values between 0.05 and 0.10 are reported in italics.

**Supplemental Table 4. Effect of sleep stabilization on other sleep and RAR metrics.**

| Outcome | Baseline weeks Mean [SD] | Manipulation week 1 Mean [SD] | Manipulation week 2 Mean [SD] | F | η2 [1] | P [2] | Q [2,3] |
|---|---|---|---|---|---|---|---|
| Sleep duration (minutes) - average | 395.06 [50.72] | 419.43 [39.25] | 409.82 [47.51] | 1.93 | 0.177 | 0.174 | 0.232 |
| Midsleep (minutes) - average | 249.3 [77.25] | 193.17 [106.9] | 195.55 [96.17] | 3.98 | 0.307 | 0.037 | 0.084 |
| Lights off time (minutes) - average | 1417.66 [61.82] | 1374.78 [118.17] | 1379.15 [99.05] | 1.42 | 0.136 | 0.268 | 0.268 |
| Get up time (minutes) - average | 485.33 [86.79] | 435.06 [96.1] | 430.87 [100.86] | 3.79 | 0.296 | **0.042** | *0.084* |

Abbreviations: RAR - rest-activity rhythms.

[1] η2 effect size benchmarks: negligible ($\eta2 \geq 0.01$), small ($0.01 \leq \eta2 < 0.06$), medium ($0.06 \leq \eta2 < 0.14$), and large ($\eta2 \geq 0.14$) partial effect sizes.

[2] P-values ≤ 0.05 are reported in bold characters; P-values between 0.05 and 0.10 are reported in italics.

[3] Q-values indicate P-values after False Discovery Rate (FDR) correction for multiple comparisons.

**Supplemental Table 5. Effect of changes (Δ) in sleep/RAR metrics on changes (Δ) in depressive and mania symptoms from pre- to post-manipulation.**

| | Δ Mood and Feelings Questionnaire (MFQ) | | | Δ Child Mania Rating Scale (CMRS)[a] | | |
|---|---|---|---|---|---|---|
| Sleep/RAR metrics | β | P[a] | Q[b,c] | β | P[a] | Q[b,c] |
| Δ Sleep duration (minutes) – variability | 0.07 | 0.789 | 0.899 | 0.10 | 0.625 | 0.847 |
| Δ Sleep duration (minutes) - average | -0.15 | 0.606 | 0.899 | -0.35 | **0.020** | *0.088* |
| Δ Midsleep (minutes) - variability | 0.07 | 0.823 | 0.899 | -0.23 | 0.243 | 0.425 |
| Δ Midsleep (minutes) - average | 0.15 | 0.641 | 0.899 | -0.39 | 0.073 | 0.170 |
| Δ CFI | -0.47 | **0.040** | 0.280 | -0.04 | 0.847 | 0.847 |
| Δ Lights off time (minutes) - average | 0.09 | 0.743 | 0.899 | -0.06 | 0.766 | 0.847 |
| Δ Get up time (minutes) - average | 0.04 | 0.899 | 0.899 | -0.45 | **0.025** | *0.088* |

Abbreviations: RAR - rest-activity rhythms; CFI - Circadian Function Index.

[a] One participant was missing CMRS data in the post-manipulation self-report.

[b] P-values ≤ 0.05 are reported in bold characters; P-values between 0.05 and 0.10 are reported in italics.

[c] Q-values indicate P-values after False Discovery Rate (FDR) correction for multiple comparisons.

**<u>SUPPLEMENTAL FIGURES</u>.**

**Supplemental Figure 1. Moderation effect of group (High-Risk versus Low-Risk) in the effects of other sleep and RAR metrics on symptom severity.**

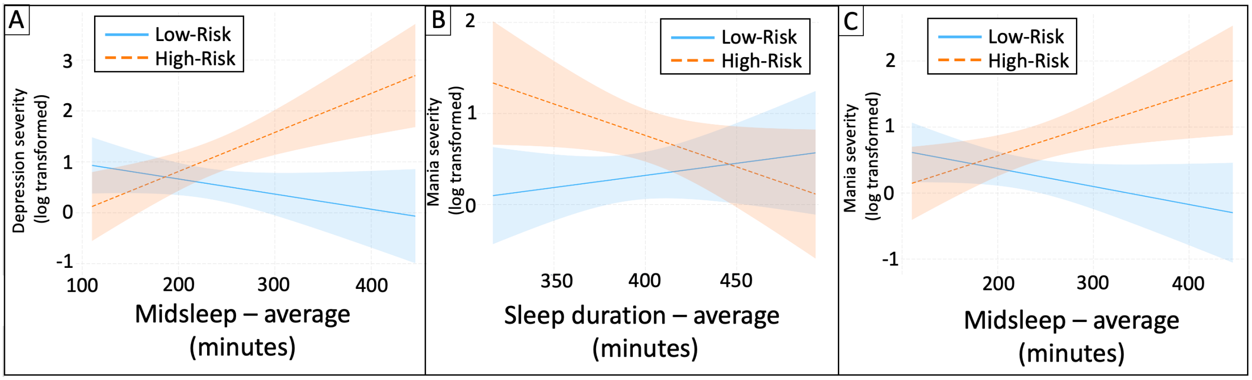


**Supplemental Figure 1 legend:** Supplemental Figure 1 shows scatter plots with the relationship between midsleep average and depression severity (KDRS; Panel A), sleep duration average and mania severity (KMRS; Panel B), and midsleep average and mania severity (KMRS; Panel C). The x-axis shows the sleep variables, and the y-axis shows the log-transformed scores of symptom severity (depression and mania). These scores were log transformed before statistical analyses due to non-normal distribution. The relationships in the High-Risk group are demonstrated in dotted regression lines while the relationships in Low-Risk groups are demonstrated in solid regression lines. The light-colored area around the regression line represents the 95% confidence interval. Only the relationships in the High-Risk group were statistically significant ($P<0.05$). Abbreviations: RAR – rest-activity rhythms; CFI – Circadian Function Index; KDRS - KSADS depression rating scale; KMRS – KSADS Mania rating scale.
